# Hyperexpanded CD4^+^ T cell clones in rheumatoid arthritis show attenuated senescence and accumulate in afflicted joints

**DOI:** 10.64898/2026.07.09.26357637

**Authors:** Phuong Nguyen, Laurin Braune, Hannah Apel, Felix Beck, Annelie K. Schierack, Roger Scholz, Lucie Loyal, Andreas Thiel, Michael Rade, Kristin Reiche, Ulrike Köhl, Tobias Hagemann, Kathrin Rothe, Ulf Wagner

## Abstract

**Objective:** Clonal hyperexpansion of CD4^+^ T cells is a characteristic feature of rheumatoid arthritis (RA). Equally large T cell clones also arise in physiological ageing or latent viral infection and adopt a replicative senescence programme – a tolerance mechanism that limits immune activation by innate-like reprogramming and proliferative arrest. We aimed to characterise the senescence programme of hyperexpanded CD4^+^ T cell clones in RA and to define their clinical associations.

**Methods:** Hyperexpanded T cell clones were characterised by single-cell RNA and T cell receptor profiling of peripheral T cells from RA patients and healthy donors. Flow cytometric validation was performed in two cross-sectional cohorts (n=15, n=45), paired blood and synovial fluid (n=20) or synovial tissue (n=18) sampling, and a non-interventional study of co-stimulatory blockade with abatacept (n=6).

**Results:** Hyperexpanded CD4^+^ T cell clones exhibited a CCR7^−^CD27^−^ phenotype and accumulated in RA joints. Their frequency correlated with disease activity and their surface profile was modulated by abatacept, suggesting susceptibility to therapeutic intervention. At the molecular level, hyperexpanded clones converged on a phenotype consistent with replicative senescence, characterised by natural killer (NK) cell-reminiscent cytotoxic reprogramming, loss of co-stimulatory molecules, and reduced translational activity. However, compared with healthy donor counterparts, hyperexpanded RA CD4^+^ T cell clones showed reduced senescence-associated cytotoxic and NK cell markers, and increased IL-7 receptor signalling, indicating attenuated senescence and preserved capacity for homeostatic proliferation.

**Conclusion:** We propose that replicative senescence insufficiently constrains hyperexpanded clones in RA, resulting in sustained antigen reactivity in autoreactive clones and perpetuation of chronic inflammation.

## Introduction

Rheumatoid arthritis (RA) is a prototypical T cell-mediated autoimmune disease whose genetic risk maps predominantly to human leukocyte antigen and T cell signalling loci^1–3^. A hallmark of the RA T cell repertoire is the accumulation of large, non-malignant CD4^+^ T cell clones, whose functional and clinical significance remains incompletely understood^4^. However, such clones also arise in healthy individuals of highly advanced age or with chronic viral infection^5,6^, leaving it unclear whether their accumulation reflects protective immunity or contributes to immunopathology.

Prior work on T cell clonal expansion, predominantly in the context of chronic viral infection and aging, identified a senescent phenotype in the largest clones, defined by the sequential loss of co-stimulatory receptors CD28 and CD27, acquisition of inhibitory receptors, telomere erosion, and proliferative arrest^7,8^. This phenotype, termed replicative senescence, arises from repeated cycles of antigen-driven clonal expansion and is considered a tolerance mechanism that limits chronic immune activation^9^. In RA, clonal CD4^+^CD28^−^ T cells expressed senescence-associated inhibitory receptors and showed telomere erosion, and were linked to autoreactivity and extra-articular manifestation^10–12^. These cells, however, do not undergo proliferative arrest and instead display a hyperproliferative phenotype, indicating that the classical senescence program is not fully operative in this context^13^.

Early studies of T cell clonal expansion relied on separate analyses of clonality and phenotype, a technical limitation that precluded insight into the transcriptional state of individual clones. Single-cell RNA sequencing with paired T cell receptor profiling now enables the simultaneous characterisation of clonal identity and gene expression at scale. Building on our prior characterisation of CD4^+^CD8α^low^ T cell clonal expansion in RA^14^, we address here the transcriptional and phenotypic identity of the largest expanding clones across both CD4^+^ and CD8^+^ T cell compartments, their disease specificity, and clinical significance. We demonstrate that the largest T cell clones, here called hyperexpanded clones, converge on a shared transcriptional program enriched for senescence-associated features in both RA patients and healthy donors. However, hyperexpanded RA CD4^+^ clones displayed a perturbed senescent phenotype relative to healthy donors, corroborating that clonal expansion in RA operates through mechanisms distinct from classical replicative senescence.

## Results

### Clonal hyperexpansion predominantly affects the CD4^+^ T cell compartment in RA

We first performed clonality analysis of 29,115 peripheral T cells in RA patients and healthy donors (HD) using an in-house single-cell RNA- and TCR-seq dataset^14^. Consistent with prior studies, most expanded T cell clones in healthy individuals derived from CD8^+^ T cells^15^. In RA, however, expanded clones were phenotypically more frequently CD4^+^ than CD8^+^ (fig.1A). The diversity of CD4^+^ T cell clones as measured by the Gini-Simpson index was reduced in RA, with no difference observed for CD8^+^ clones (fig. 1B, fig. S1A).

**Figure 1:**
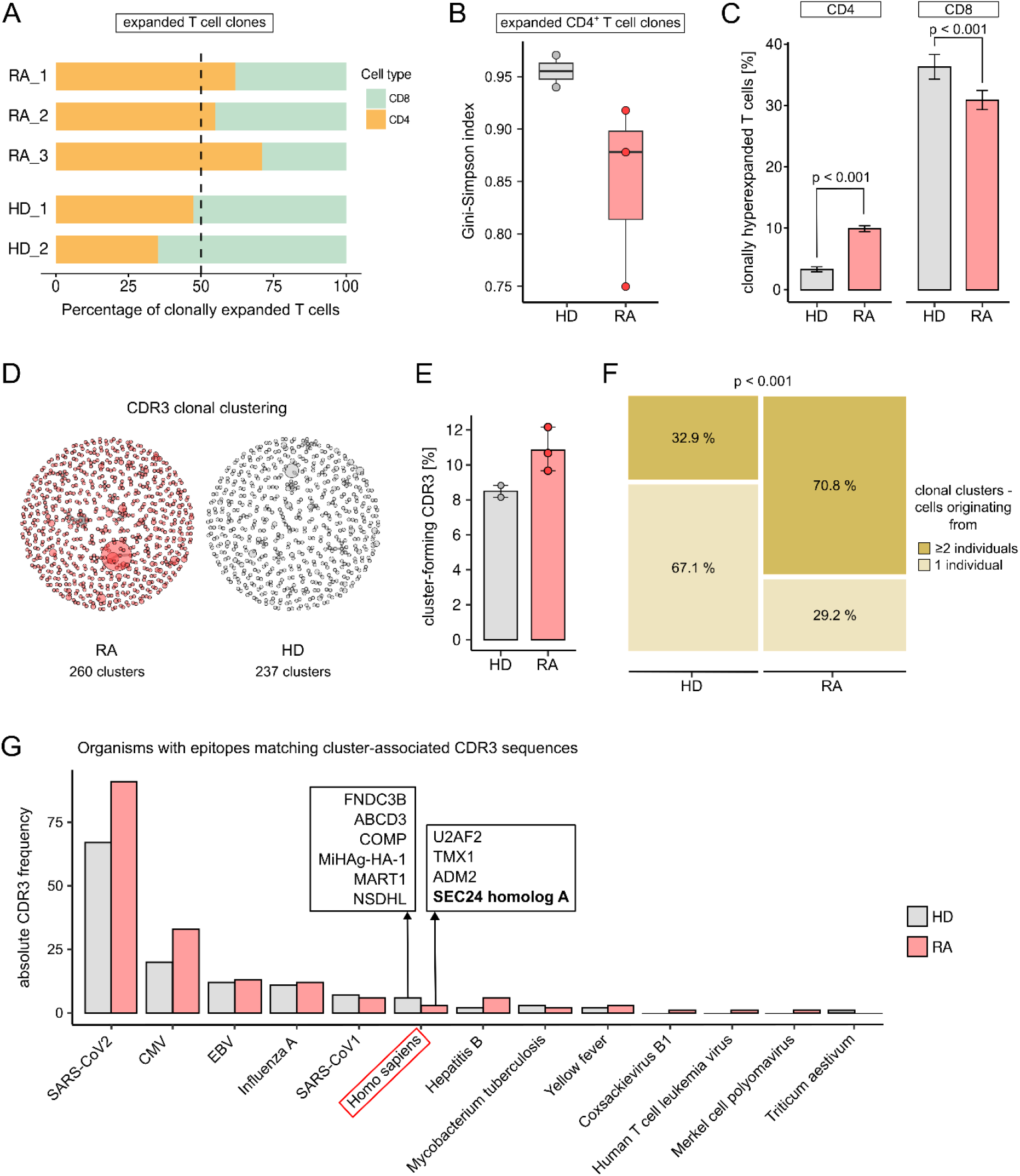
Amplified clonal hyperexpansion is likely antigen-driven in RA CD4^+^ T cells. **A)** The distribution of CD4^+^ and CD8^+^ T cells within all expanded T cell clones for each individual. **B)** Gini-Simpson index reflecting the TCR diversity of expanded CD4^+^ T cell clones in RA and HD. **C)** Proportion of cells belonging to a hyperexpanded T cell clone (≥ 10 T cells sharing the same α/β-TCR sequence) in the CD4^+^ and CD8^+^ T cell compartments. Error bars indicate 95% confidence interval. **D, E)** Similar CDR3 sequences of the TCR β-chain of CD4^+^ T cells are grouped into clonal clusters based on a low Levenshtein edit distance. Network plot **(D)** depicting the results of clonal clustering. Each node or circle is a clone with the same CDR3 sequence, its size corresponding to the number of cells expressing that CDR3 sequence. Clones with similar CDR3 sequences belong to one clonal cluster and are connected by grey edges. **E)** Proportion of CDR3 sequences that are part of a clonal cluster in each individual, showing CDR3 sequences of RA samples to cluster together more frequently than in HD. **F)** Number of individuals each clonal cluster originates from. Indicated p-value for Chi-square test comparing the proportion of clonal clusters originating from one or multiple individuals between RA and HD. **G)** Matching epitopes for each cluster-associated CDR3 sequence were searched for in a public database. The organisms those epitopes belonged to are plotted against the number of matching cluster-forming CDR3 sequences. Six and four autoantigenic epitopes are found for HD and RA CDR3 sequences, respectively. scRNA/TCR-seq, single-cell RNA and T cell receptor sequencing. HD, healthy donor. RA, rheumatoid arthritis. CDR3, complementarity-determining region 3.

This CD4^+^-skewed pattern in RA was most pronounced among exceptionally large clones, hereafter termed hyperexpanded clones (≥10 of all analysed cells sharing an identical paired α/β-TCR sequence). Hyperexpanded clones were three times more abundant in the CD4^+^ compartment of RA patients than HD (9.9% vs. 3.3%, fig. 1C), whereas their proportion within the CD8^+^ compartment was slightly more prominent in HD (30.8% vs. 36.2%, fig. 1C). Small, conventional clones (>1 and <10 cells with identical α/β-TCR sequence), in contrast, were not more frequent in RA (fig S1B). This suggests that clonal hyperexpansion of CD4^+^ T cells is RA specific, while large CD8^+^ clones are a normal feature of peripheral T cell homeostasis.

### CD4^+^ T cell clonal expansion is likely to be antigen-driven in RA

We next assessed TCR sequence similarity among CD4^+^ T cells across samples. No exact clonotype overlap was detected between individuals, nor was significant preferential VDJ gene usage observed (fig. S1C, S1D). Since antigen recognition is primarily determined by the complementarity-determining region 3 (CDR3) of the TCR, highly similar CDR3 sequences likely share epitope specificity. We therefore grouped highly similar CDR3 sequences of CD4^+^ TCR β-chains into clonal clusters predicted to recognise related epitopes. Despite the absence of clonotype overlap, RA CDR3 sequences were more similar to one another than in HD, reflected by more clonal clusters (260 vs. 237, fig. 1D), a higher proportion of cluster-forming CDR3 sequences among all sequences analysed (mean 10.8% vs. 8.5%, fig. 1E), and larger cluster size (2.9±2.2 vs. 2.6±1.5 sequences/cluster). The majority of RA clusters contained CDR3 sequences from two or more patients, whereas HD clusters mainly comprised sequences from a single individual (Chi-square p < 0.001, fig. 1F), underscoring the inter-individual sequence similarity in RA.

Next, we identified potential target antigens by querying cluster-forming CDR3 sequences against the IEDB epitope database. Matches were obtained for 15.9% and 13.3% of sequences in RA and HD, respectively, dominated by SARS-CoV-2, CMV, EBV, and Influenza A, reflecting the known viral bias of public epitope databases (fig. 1G). No significant between-group difference was detected (table S1, S2), arguing against differential viral exposure as a driver of CD4^+^ clonal expansion in RA. Autoantigen matches were modest in both groups (3 autoreactive CDR3 sequences mapping to 4 epitopes in RA; 6 sequences mapping to 6 epitopes in HD), consistent with expected background autoreactivity in healthy individuals^16^. Although the cohort was not powered to identify disease-specific autoantigens, we noted one autoantigen that matched a CDR3 sequence shared across all RA samples: SEC24 homolog A (SEC24A), a COPII vesicle coat component required for endoplasmic reticulum export^17^, which has not been previously reported in RA.

Taken together, our findings identify clonal hyperexpansion of CD4^+^ T cells as a distinct feature of RA. The cross-patient CDR3 sequence similarity, which is insufficiently explained by viral exposure alone, points toward shared, MHC class II-restricted autoantigen recognition as a driver for CD4^+^ clonal hyperexpansion in RA.

### Hyperexpanded T cell clones adopt a senescent phenotype and lose CD27 in RA patients and healthy donors

We next characterised the transcriptional changes associated with clonal hyperexpansion. Hyperexpanded clones in CD4^+^ and CD8^+^ compartments shared a largely overlapping transcriptional program, with 1533 differentially expressed genes (DEGs, FDR <0.05) regulated simultaneously in both T cell subsets (fig. 2A, B). In contrast, small conventional clones retained the distinct helper and cytotoxic programs expected of CD4^+^ and CD8^+^ T cells (fig. 2C). The shared hyperexpansion program was defined by four features: upregulation of cytotoxicity-related genes (GZMB, GNLY, FGFBP2) alongside innate-like NK markers (NKG2D/KLRK1, KLRF1, CD56/NCAM1); a late-activated, migratory phenotype (downregulation of IL2RA/CD25; upregulation of HLA class II, CX3CR1, CCL4, CCL5); downregulation of naïve and central memory markers (MAL, SELL, CCR7, CD27); and concomitant upregulation of transcription factors associated with terminal differentiation (HOPX, EOMES, ZEB2) (fig. 2B, C). Collectively, this transcriptional profile aligns closely with signatures of T cell replicative senescence^18^, supported by gene set enrichment showing upregulated NK-cell cytotoxicity/degranulation pathways and downregulated translational activity – a hallmark of proliferation arrest in senescent cells (fig. 2D).

**Figure 2:**
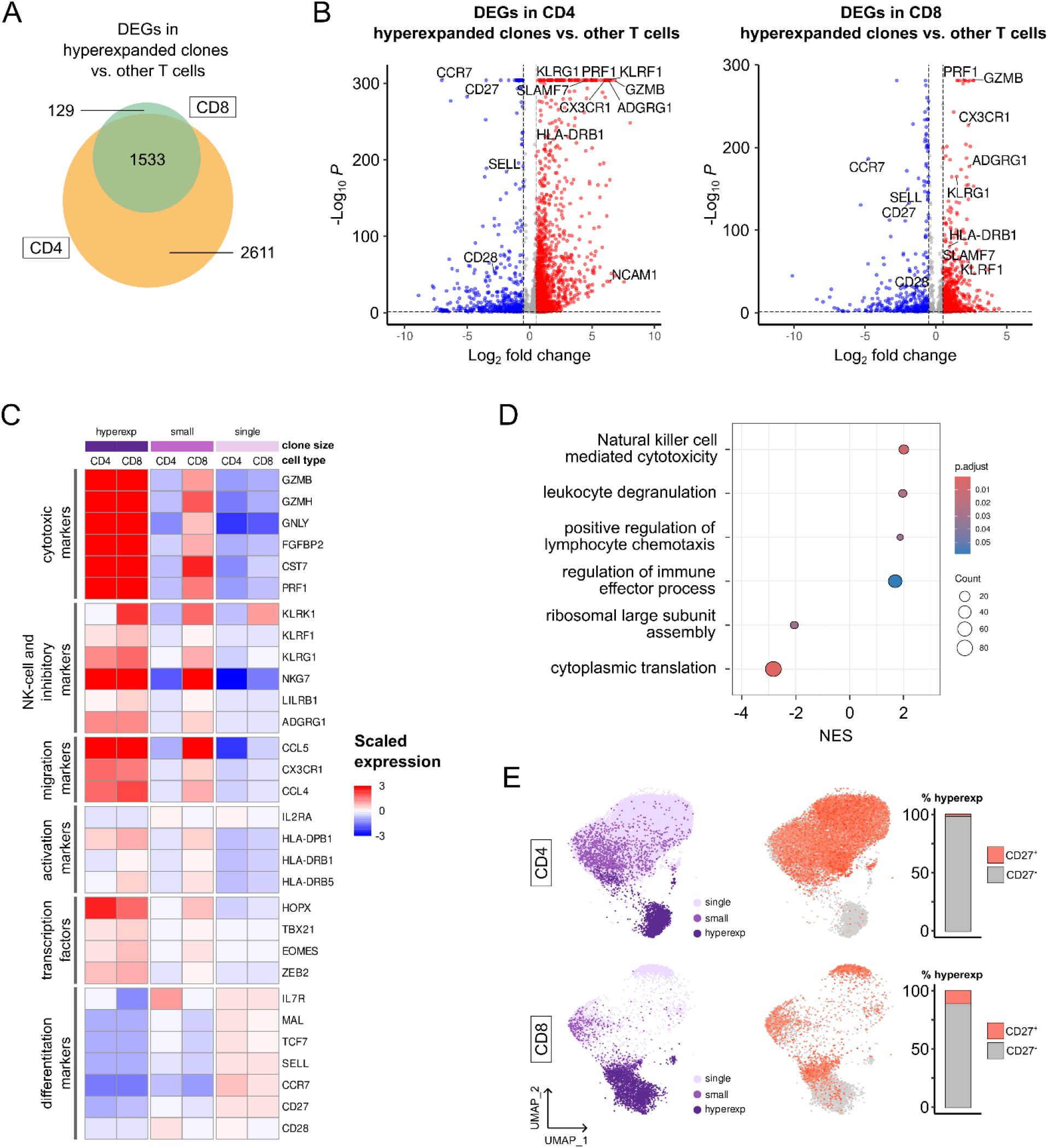
T cell clonal hyperexpansion features signatures of replicative senescence and is marked by CD27 loss. **A)** Differential gene expression analysis revealed 1533 genes to be commonly differentially regulated in CD4^+^ and CD8^+^ hyperexpanded T cell clones compared to non-clonally hyperexpanded T cells. In contrast, 2611 genes are exclusively differentially expressed in CD4^+^ hyperexpanded clones and 129 genes exclusively in CD8^+^ hyperexpanded clones compared other CD4^+^ and CD8^+^ T cells, respectively **B)** The overlapping DEGs are similarly up- and downregulated in CD4^+^ and CD8^+^ hyperexpanded T cell clones compared to non-hyperexpanded clones. **C)** Heatmap showing the scaled expression of selected DEGs that characterize clonal hyperexpansion, separated by clone size and affiliation with the CD4^+^ or CD8^+^ T cell compartment. Hyperexpanded clones are defined as having ≥10, small clones as having at least 2 but <10 T cells sharing an identical paired α/β-TCR sequence. Singletons have a unique TCR sequence. **D)** Gene set enrichment analysis of differentially expressed genes in clonally hyperexpanded CD4^+^ T cells showing upregulation of cytotoxicity and degranulation pathways, chemotaxis and effector processes, and downregulation of translational pathways. **E)** UMAPs showing the distribution of singletons, small clones and hyperexpanded clones (left-hand side) and corresponding expression of CD27 (right-hand side). UMAPs in the upper row show CD4^+^ T cells, in the lower row CD8^+^ T cells. DEG, differentially expressed gene. NES, normalized enrichment score. Hyperexp, hyperexpanded clones. UMAP, uniform manifold approximation and projection

Among DEGs encoding surface proteins, CCR7 and CD27 were consistently downregulated in hyperexpanded CD4^+^ and CD8^+^ clones (fig. 2E, fig. S2A). Their expression correlated inversely with clone size (R = −0.34 and −0.39, p < 0.001, fig. S2B). CD27 loss was especially characteristic of hyperexpanded CD4^+^ clones, with only 2.1% retaining CD27 expression (fig. 2E). Consistent with previous literature, we proceeded to identify clonally hyperexpanded cells by the CCR7^−^CD27^−^ phenotype and small conventional effector memory clones as CCR7^−^CD27^+^ for flow cytometry analysis^19^ (fig. S3A).

In summary, hyperexpanded T cell clones exhibit a senescent transcriptional program of innate-like cytotoxicity, late activation, migratory capacity, and translational downregulation, with CCR7 and CD27 loss as reliable surrogate markers.

### Clonally hyperexpanded CD4^+^ T cells are not fully committed to senescence in RA

We next asked whether the senescent transcriptional program differs between RA and healthy hyperexpanded clones. Differential gene expression analysis identified 339 DEGs (FDR <0.05) between RA and HD hyperexpanded CD4^+^ T cell clones (fig. 3A). Several hyperexpansion markers, including PRF1 and CD8A, were more strongly upregulated in RA. Gene ontology enrichment analysis further distinguished RA hyperexpanded clones by type I interferon pathway genes (JAK1, IFITM3, IRF1), IL-7 signalling components (PDIA3, P4HB), and tumour necrosis factor-mediated signalling (TNFRSF1B, GSTP1, CARD16; fig. 3A, B). Conversely, apoptosis genes, response to reactive oxygen species, and ribosomal genes were downregulated relative to HD, suggesting a long-lived, translationally quiescent state (fig. 3A, B).

**Figure 3:**
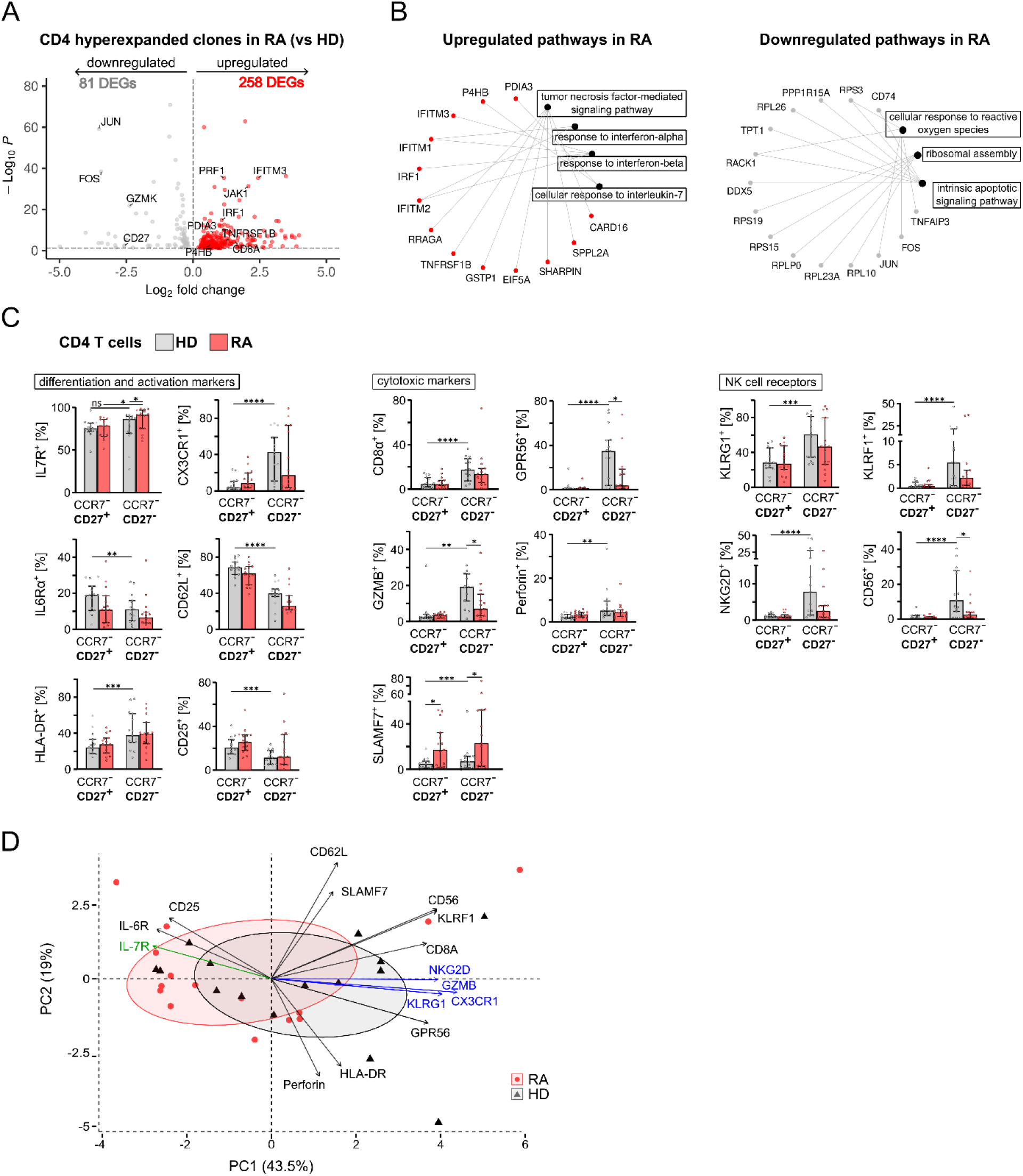
Clonally hyperexpanded CD4^+^ T cells in RA are not fully committed to senescence. **A)** Differentially expressed genes between clonally hyperexpanded CD4^+^ T cells in RA and HD. **B)** Pathway analysis showing upregulated and downregulated pathways (black nodes) and corresponding marker genes (red nodes for upregulated, grey nodes for downregulated genes in RA compared to HD). **C)** Marker expression in clonally hyperexpanded (CCR7^-^CD27^-^) and conventional effector memory clones (CCR7^-^CD27^+^) in RA and HD CD4^+^ T cells (n = 15 each). **D)** Marker expression in clonally hyperexpanded CD4^+^ T cells were subjected to principal component analysis. The first two principal component are plotted, together explaining 62.5% of the total variance. Individual data points are marked as red circles (RA) and black triangles (HD). Vectors indicate marker loadings for each principal component. The strongest senescence-associated markers are coloured blue, while IL-7 receptor is coloured green to highlight their opposing contribution to PC1. Data in bar charts are shown as median ± interquartile range. Ns = non-significant, \**p* ≤ 0.05, \*\**p* ≤ 0.01, \*\*\**p* ≤ 0.001, \*\*\*\**p* ≤ 0.0001 for Mann-Whitney-U test for unpaired data. RA, rheumatoid arthritis. HD, healthy donors. PC, principal component.

In a pilot validation cohort (n=15 RA, 15 HD), flow cytometry confirmed that clonally hyperexpanded CCR7^−^CD27^−^CD4^+^ T cells showed higher expression of NK receptors, cytotoxic markers, and migration marker CX3CR1 than conventional CCR7^−^CD27^+^CD4^+^ clones in both RA and HD, consistent with the senescent phenotype at the RNA level (fig. 3C). The late activation marker HLA-DR was upregulated, while early markers CD25/IL2-RA and IL-6 receptor (progressively lost upon activation) were downregulated in hyperexpanded versus conventional clones, indicating hyperexpanded clones might have passed peak activation.

When comparing HD and RA CCR7^−^CD27^−^CD4^+^ T cells, we found cytotoxicity and NK-cell markers ADGRG1/GPR56, GZMB and CD56 to be more highly expressed in HD, pointing to a deeper senescent commitment in healthy individuals (fig. 3C). On the other hand, IL-7 receptor was upregulated in RA (fig. 3B, C), corroborating the transcriptomic data and indicating retained capacity for IL-7-mediated homeostatic proliferation.

Dimensionality reduction of the full flow cytometry marker panel reinforced this picture. Principal component analysis (62.5% variance explained across two components, fig. 3D) revealed that the first component, driven by correlating senescent markers NKG2D, KLRG1, CX3CR1, and GZMB, segregated HD from RA: HD samples clustered toward higher senescent marker expression, while RA samples were displaced toward IL-7 receptor expression. Hierarchical clustering yielded a consistent result, with senescence clusters enriched for HD (61.5% vs. 38.5% RA) and non-senescence clusters enriched for RA (58.8% vs. 41.2% HD; fig. S4). Together, these findings indicate that hyperexpanded CD4^+^ clones in HD adopt a more senescent, terminally differentiated phenotype, whereas those in RA retain a less senescent, cytokine-responsive state.

### Hyperexpanded CD4^+^ T cell clones accumulate in RA joints and express PD-1

We next examined whether clonally hyperexpanded CD4^+^ T cells preferentially traffic to the inflamed joint. Peripheral blood paired with synovial tissue (n=18) or synovial fluid (n=20) were obtained from RA patients and from 5 patients with osteoarthritis (OA) as controls. CCR7^−^CD27^−^CD4^+^ clonally hyperexpanded T cells were significantly enriched in both synovial compartments relative to peripheral blood in RA patients (fig. 4A), while no significant difference between synovial fluid and synovial tissue was discernible (30.2% vs. 23.0% of CD4^+^ T cells, p=0.13). Frequencies of hyperexpanded clone cells in blood correlated with synovial fluid frequency (r=0.60, p=0.005; fig. 4B) but not with synovial tissue frequency, suggesting active cellular exchange between blood and synovial fluid.

**Figure 4:**
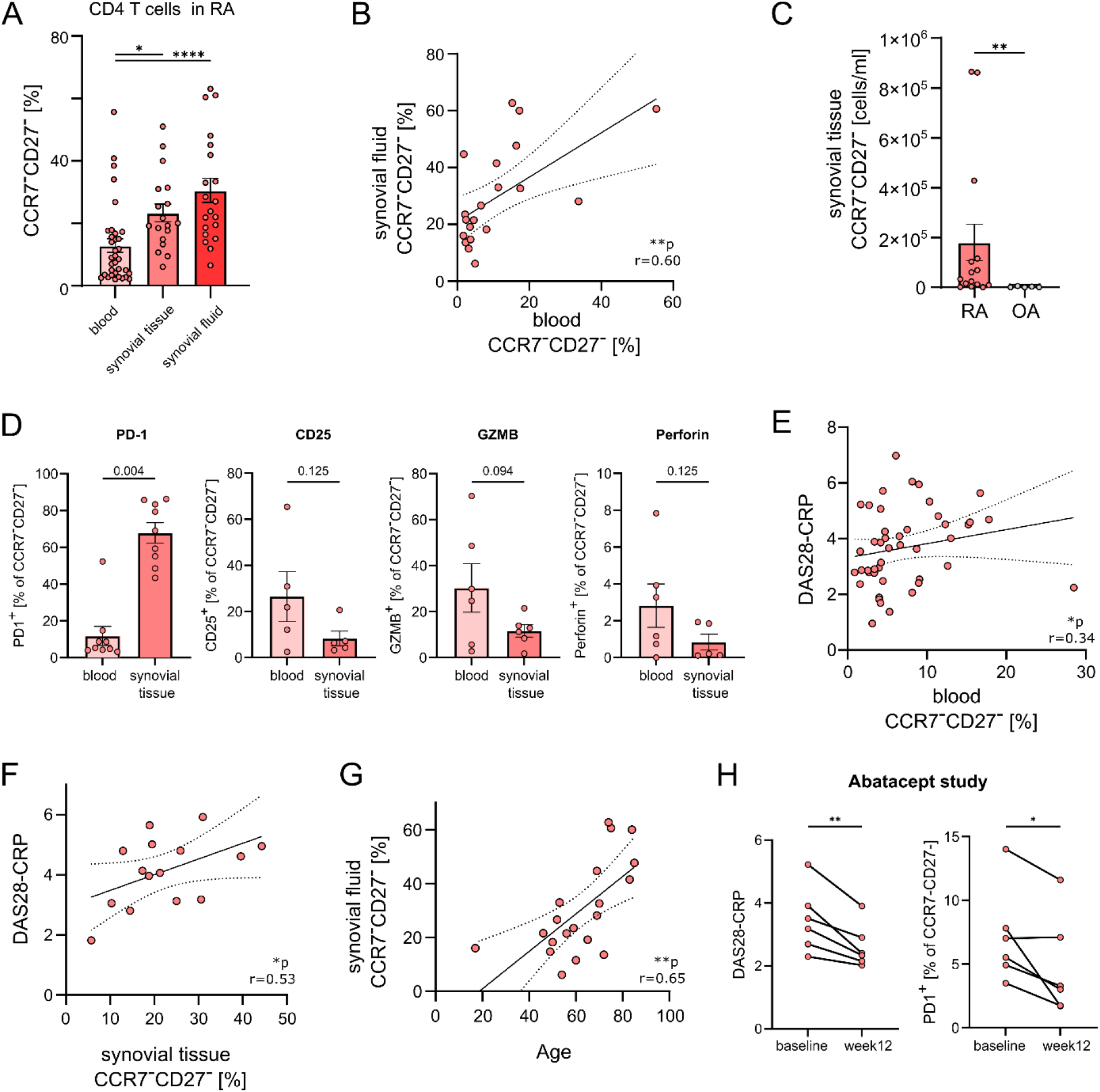
Hyperexpanded CD4^+^ T cell clones accumulate in RA joints and are associated with systemic and local inflammation. **A)** The proportion of hyperexpanded clones among CD4^+^ T cells in different anatomical compartments in RA patients (paired blood-tissue samples n=18, paired blood-fluid samples n=20). **B)** Positive correlation between hyperexpanded CD4^+^ clones in synovial fluid and peripheral blood in RA samples. **C)** Absolute counts of hyperexpanded CD4^+^ T cell clones in RA vs. OA synovial tissue (n=15 and 5). **D)** Expression of PD-1 (n=9), CD25 (n=5), GZMB (n=6), Perforin (n=6) on hyperexpanded CD4^+^ clones in RA blood vs. synovial tissue. **E, F)** Positive correlation of RA disease activity measured by DAS28-CRP with hyperexpanded CD4^+^ clone frequency in synovial tissue (n=15, **F**) and in peripheral blood of an independent cohort (n=45, **E**). **G)** Correlation of hyperexpanded CD4^+^ clones in synovial fluid with age (n=20). **H)** Reduced disease activity DAS28-CRP and downregulated expression of PD-1 on hyperexpanded CD4^+^ T cell clones in blood of RA patients 12 weeks after treatment initiation with abatacept (n=6). Data in bar charts are displayed as mean ± standard error mean. \**p* ≤ 0.05, \*\**p* ≤ 0.01, \*\*\**p* ≤ 0.001, \*\*\*\**p* ≤ 0.0001 for paired Wilcoxon signed-ranked (A, D) or unpaired Mann-Whitney-U test (C), paired t-test (H), as well as Spearman’s correlation (B, E-G). RA, rheumatoid arthritis. OA, osteoarthritis. DAS28-CRP, 28-joint Disease Activity Score with C-reactive protein.

Absolute counts of clonally hyperexpanded CD4^+^ T cells in RA synovial tissue substantially exceeded OA (mean 1.7×10^5^ vs. 0.02×10^5^ cells/ml, p=0.005; fig. 4C), indicating that joint enrichment reflects active inflammatory infiltration in RA.

We then characterised the phenotype of hyperexpanded clones in the synovial microenvironment. Among the markers assessed, PD-1 was most strikingly upregulated on synovial tissue hyperexpanded clones relative to blood (mean 67.8% vs. 11.8% of CCR7^−^CD27^−^CD4^+^ T cells, p=0.004, fig. 4D), consistent with prior reports of PD-1^high^ peripheral T helper cell accumulation in RA joints^20,21^. Downregulation of CD25, GZMB, and perforin in synovial tissue was directionally consistent but did not reach statistical significance (fig. 4D).

### Disease activity correlates with the frequency of hyperexpanded CD4^+^ T cell clones in both synovial tissue and peripheral blood

Given the accumulation of hyperexpanded clones in afflicted joints, we examined further clinical associations in a cross-sectional cohort expanded to 45 RA patients with varying disease duration. Disease activity was quantified by the 28-joint Disease Activity Score with C-reactive protein (DAS28-CRP; mean ± SD = 3.7 ± 1.4; table S3A). Peripheral blood CCR7^−^CD27^−^CD4^+^ T cell frequency correlated with DAS28-CRP (r = 0.34, p = 0.021; fig. 4E), with comparable correlations for SDAI and serum CRP (fig. S5A, B), but not disease duration (fig. S5C).

In the synovial cohort, the positive correlation between synovial tissue CCR7^−^CD27^−^CD4^+^ T cell frequency and DAS28-CRP was stronger than in blood (r = 0.53, p = 0.045; fig. 4F). CCR7^−^CD27^−^CD4^+^ T cell frequency also correlated with age across all compartments, strongest in synovial fluid (r = 0.65, p = 0.002; fig. 4G, fig. S5D, E). A multivariable regression model with synovial tissue CCR7^−^CD27^−^CD4^+^ T cell frequency as the outcome identified both disease activity and age as independent predictors (DAS28-CRP: β-coefficient = 4.22, p = 0.019; age: β-coefficient = 0.56, p = 0.002; table S4).

### Hyperexpanded CD4^+^ T cell clones respond to efficacious T cell co-stimulation blockade

Finally, in order to investigate the link between hyperexpanded CD4^+^ clones and disease activity *in vivo*, a non-interventional clinical study was initiated in RA patients previously not treated with biological or targeted-synthetic disease-modifying antirheumatic drugs, all of whom required a treatment escalation due to inadequate disease control (DAS28-CRP 4.06 ± 0.82, mean ± SD). Patients were started on abatacept, a selective T cell co-stimulatory blocker, and clinical response, disease activity, and hyperexpanded CD4^+^ clones were reassessed after 12 weeks.”

All six patients achieved good clinical response (DAS28-CRP = 2.62 ± 0.70 at week 12, fig. 4H). The frequency of circulating CCR7^−^CD27^−^CD4^+^ T cells did not change, but PD-1^+^ cells within this population declined significantly after 12 weeks (mean 7.1% versus 4.7%, p=0.035; fig. 4H), consistent with a role for T cell co-stimulation in maintaining PD-1 expression on hyperexpanded CD4^+^ T cell clones.

Together, our data indicate that CCR7^−^CD27^−^CD4^+^ hyperexpanded clones migrate to RA joints, upregulate PD-1 expression in the synovial microenvironment, and contribute to both local and systemic inflammation, likely in a co-stimulation-dependent manner.

## Discussion

Perturbation of the T cell repertoire is a characteristic feature of RA, underscoring the central pathophysiological role of T cells in the disease. Previous work reported marked T cell repertoire contraction in RA, with median TCR β-chain frequencies up to 10-fold higher in CD4+ T cells than in healthy individuals^4^. Our single-cell data corroborate this skew by showing that hyperexpanded RA CD4^+^ T cell clones were three times more abundant than in healthy controls (fig. 1C).

One possible mechanism underlying CD4^+^ T cell oligoclonality in RA is persistent antigenic pressure. This aligns with the strongest genetic risk factor in RA: certain HLA-DRB1 alleles encode MHC class II molecules that preferentially present autoantigens to CD4^+^ T cells, thereby generating chronic antigenic stimulation that drives clonal expansion^2^. Although epitope identification remains technically demanding and the RA autoantigen repertoire beyond citrullinated peptides is incompletely defined, evidence for shared antigen recognition is accumulating. Recent work using single-cell TCR sequencing reported a TRBV20-1 gene usage bias in an anti-citrullinated-protein antibody-positive cohort^22^, and increased TCR clustering across patients in clonal T cells from lung, joint and blood in another^23^. Consistent with this, our data showed elevated inter-individual CDR3 β-chain sequence similarity in RA relative to healthy controls (fig. 1D–F), suggesting recognition of shared MHC class II-restricted epitopes.

Antigen-independent mechanisms could also contribute to clonal hyperexpansion. The RA T cell compartment is susceptible to lymphopenia, possibly due to deficient thymic output or deficiency of the homeostatic cytokine IL-7^24,25^. Upregulation of the IL-7 receptor in hyperexpanded CD4^+^ clones (fig. 3C) might reflect a compensatory response, and rising IL-7 concentrations during remission periods or after successful therapy could, in turn, result in clonal outgrowth^26^. Furthermore, oligoclonality is already present in naïve RA T cells, which have not encountered their cognate antigen, further supporting an antigen-independent contribution^4^. Clonal hyperexpansion in RA thus most likely reflects combined effects of antigen-driven and antigen-independent proliferation.

At the molecular level, hyperexpanded T cell clones converged on a phenotype consistent with replicative senescence (fig. 3B-C). Replicative senescence is a tolerance mechanism in which repeated antigen-driven proliferation leads to proliferative arrest and replacement of antigen-specific function with NK cell-like, non-specific effector activity^7,9^. This programme limits pathological immune over-activation and reduces the risk of malignant transformation from uncontrolled lymphocyte proliferation. In healthy individuals, this is most prominent in T cell clones driven by persistent cytomegalovirus infection^5,27^. In autoimmune diseases, an analogous mechanism appears to be engaged in response to persistent autoantigen stimulation: a senescent-like phenotype has been described for CD4^+^ T cell clones in RA^21,28^, and for pathogenic, disease-specific CD4^+^ T cells in Guillain-Barré syndrome and type 1 diabetes^29,30^. These data suggest replicative senescence is a broadly operative, if not invariably sufficient, response to chronic T cell stimulation.

Hyperexpanded CD4^+^ clones in RA displayed an attenuated senescence phenotype relative to healthy controls, suggesting replicative senescence is dysregulated in disease. RA clones expressed lower cytotoxic effector molecules and NK-lineage receptors, and higher IL-7 receptor expression (fig. 3C) – a pattern fitting antigen-experienced memory T cells capable of homeostatic proliferation. Consistent with this, previous work described decreased cytotoxic degranulation^27^ and impaired cell-cycle arrest in RA senescent CD4^+^ T cells, resulting in hyperproliferation and tissue invasiveness^13^. We further identified upregulation of interferon type I and tumour necrosis factor response programmes in RA (fig. 3B), consistent with ongoing inflammatory activation. Together, these findings suggest an attenuated senescent phenotype and thus retained capacity for antigen-specific immune activation in hyperexpanded RA CD4^+^ clones. If these clones are autoreactive, failure to suppress them through replicative senescence may perpetuate chronic autoimmunity and inflammation.

Underscoring their pathogenic role, hyperexpanded CD4^+^ T cell clones accumulated in synovial compartments relative to peripheral blood in RA (fig. 4A). Blood and synovial fluid hyperexpanded clone frequencies showed a positive correlation (fig. 4B), indicating active trafficking, retention, and possibly proliferation within the inflamed joint. CX3CR1 expression on hyperexpanded clones (fig. 2C) provide a plausible recruitment mechanism, as its ligand CX3CL1 is upregulated on synovial fibroblasts and acts as a proinflammatory co-stimulator of T cell activation under inflammatory conditions in RA^31,32^.

Hyperexpanded CD4^+^ clone frequency in synovial tissue and blood correlated with DAS28-CRP (fig. 4E-F), linking accumulation of this population to disease activity. In the synovial microenvironment, these cells displayed elevated PD-1 and reduced GZMB and perforin relative to blood (fig. 4D), consistent with the T peripheral helper (Tph) phenotype previously described to clonally expand in RA synovium^20,21^. Tph cells are the main PD-1-expressing population in RA synovium^33^, show a CCR7^−^CD27^−^ phenotype and sustain local B cell-dependent inflammation through antigen-dependent helper function^20^ – a role mechanistically distinct from the non-specific cytotoxicity of terminally differentiated, senescent T cells. This finding extends the dysregulated senescence argument to the inflamed joint: rather than progressing toward senescent arrest, clonally hyperexpanded cells acquire a helper programme that actively sustains local inflammation. Co-stimulatory signalling may help maintain this Tph-like state, as PD-1 expression on CCR7^−^CD27^−^CD4^+^ T cells declined after 12 weeks of abatacept in our pilot substudy (fig. 4H).

Several limitations merit consideration. The scRNA-seq cohort was limited in size, constraining statistical power at the transcriptional level. However, key findings were replicated in a larger, independent protein-level cohorts, supporting generalisability. Mechanistic evidence of the effector capacity of hyperexpanded CD4^+^ clones and of the antigen specificities driving their expansion in RA remains to be generated. This work nonetheless provides a comprehensive single-cell molecular characterisation of hyperexpanded CD4^+^ T cell clones in RA, establishes their clinical relevance through association with disease activity and response to co-stimulatory blockade, and lays groundwork for mechanistic and interventional studies addressing whether selective targeting of these clones could attenuate disease.

## Methods

### Patient recruitment

Peripheral blood was collected from 45 RA patients attending the Rheumatology Clinic of the University of Leipzig Medical Center, all fulfilling the 2010 ACR/EULAR classification criteria. Age-matched healthy donors (n=44) served as controls. An independent cohort of RA patients (n=34) and osteoarthritis patients (n=5) undergoing joint surgery was recruited from the Collm Clinic for Orthopaedic Surgery in Oschatz, Germany. From this cohort, matched samples of blood and synovial fluid were available for 20 patients, synovial tissue for 18 patients, and some patients contributed to both compartments. For the abatacept study, six RA patients with inadequate response to methotrexate initiated subcutaneous abatacept (125 mg/week). Blood was collected at baseline and at week 12. All participants provided written informed consent. The study was approved by the ethics committee of Leipzig University. Patient characteristics for each cohort are summarised in table S3.

### Patient and Public Involvement

Patients or the public were not involved in the design, conduct, and reporting of this research.

### Peripheral blood and synovial samples processing

Peripheral blood mononuclear cells and synovial fluid mononuclear cells were isolated by Ficoll density gradient centrifugation. Synovial membrane tissue was enzymatically digested using 30 mg collagenase, 10 mg hyaluronidase, and 1 mg DNase per 10ml enzyme suspension and incubated for 90 min at 37 °C in 5% CO_2_. Cell suspension was filtered through gauze followed by 70 μm cell strainers to remove tissue debris. Viable cell counts were determined by haemocytometer counting using trypan blue exclusion, and the tissue volume (ml) was recorded.

### Flow cytometry

Cryopreserved cells (10% DMSO/fetal bovine serum) were thawed or fresh cells used directly for flow cytometry. Non-specific binding was blocked using heat-inactivated (56°C, 20 minutes) human Fcγ-binding serum. Cells were stained with fixable viability dye and fluorochrome-conjugated antibodies (table S5), then incubated at 4°C (surface antigens: 10 minutes, intracellular antigens: 30 minutes). Intracellular staining followed standard fixation/permeabilization protocols, using BD Cytofix/Cytoperm™ Fixation/Permeabilization Kit (BD Biosciences). Data were acquired on a BD LSRFortessa™ and analysed using FlowJo v10 (gating strategy in fig. S3). Principal component analysis of surface marker expression was performed in R v4.3.2 using the package FactoMineR v2.14 and factoextra v2.0.0^34^, with function PCA() and argument scale.unit = TRUE, and a biplot created using fviz_pca_biplot() with argument ellipse.level = 0.5.

For synovial tissue cells, 1×10^6^ viable cells per sample were stained and all events acquired until near-exhaustion of the tube. Absolute counts of CCR7^−^CD27^−^CD4^+^ T cells in synovial tissue were determined using following formula:

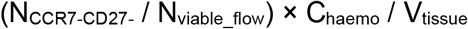

where

N_CCR7-CD27-_: is the number of CCR7^−^CD27^−^ CD4^+^ T cell events acquired by flow cytometry,

N_viable_flow_: is the total number of viable events acquired in the same tube,

C_haemo_: is the total viable cell count of the tissue digest as determined by haemocytometer,

V_tissue_: is the tissue volume in ml.

### Single-cell sequencing analysis

Single-cell RNA and TCR sequencing, preprocessing and CD4^+^ and CD8^+^ T cell annotation were described elsewhere^14^. CD4^+^ and CD8^+^ T cells were analysed separately. For each lineage, cells were classified according to the T cell clone they belong to: hyperexpanded clones (≥10 cells sharing an identical TCRα/β pair within one individual), small conventional clones (>1 and <10 cells), or singletons (1 cell).

The package scRepertoire v2.2.1 was used for all TCR analysis^35^. Clonality and diversity metrics were calculated using clonalDiversity() with arguments cloneCall = “strict” and n.boots = 500. For CDR3 analysis, amino acid sequences were obtained for all CD4^+^ T cell TCR β-chains, then randomly downsampled to equal numbers across RA and HD (n=6929). The Levenshtein distance and clustering of similar CDR3 amino acid sequences was performed using clonalCluster() with arguments chain = “TRB”, sequence = “aa”, threshold = 0.85. Clustering results were visualised as a network plot with node size correlating to number of cells sharing a CDR3 sequence, and edges connecting highly similar CDR3 sequences to a clonal cluster.

The CDR3 sequences that formed a clonal cluster were extracted separately for the RA and HD cohorts and pasted into the TCRMatch tool of the IEDB database using the default settings (https://tools.iedb.org/tcrmatch). Predicted epitope specificity and the organism it belongs to is provided for each matched CDR3 sequence in table S1.

Differential gene expression analysis was based on FindMarkers() with default settings in Seurat v5.1.0^36^. HLA genes were excluded from between-group differential expression analyses due to the potential confounding effect of inter-individual allele variation. Pathway analysis was performed using the packages clusterProfiler v4.10.1 and enrichplot v1.22.0^37^. Gene set enrichment analysis was based on Gene Ontology Biological Processes obtained from the R package org.Hs.eg.db v.3.18.0 and was applied for differentially expressed genes (FDR <0.05) in clonally hyperexpanded versus other CD4^+^ T cells using gseGO() with arguments minGSSize = 3, maxGSSize = 1000, pvalueCutoff = 0.1, pAdjustMethod = “BH”. Pathway overrepresentation analysis was performed for CD4^+^ hyperexpanded clones in RA vs. HD and was also based on Gene Ontology Biological Processes. Function enrichGO() was used for upregulated and downregulated genes, with pvalueCutoff = 0.1 and pAdjustMethod = “BH”.

All downstream analysis and visualization utilized R v4.3.2, Seurat v5.1.0 and ggplot v3.4.4.

### Statistical analysis

Continuous variables between two independent groups (e.g. RA vs. OA, RA vs. HD) were compared using the Mann–Whitney U test (non-normally distributed) or Welch’s t-test (normally distributed). Within-patient comparisons across compartments (blood vs. synovial tissue, blood vs. synovial fluid) used the Wilcoxon signed-rank test. Proportions between groups (hyperexpanded clones in RA vs. HD, clonal clusters from single vs. multiple individuals) were compared by Chi-squared test. Correlations were assessed using Spearman’s rank correlation coefficient. Longitudinal comparisons (abatacept study, baseline vs. week 12) used the Wilcoxon signed-rank test. All tests were two-tailed; p < 0.05 was considered significant. Analyses used R v4.3.2 and GraphPad Prism v11.0.0.

## Data Availability

The single-cell sequencing data generated in this study are available from the corresponding author upon reasonable request.

## Acknowledgements

We thank Dr. Caroline Schmidt for critical reading of the manuscript and helpful discussions. We thank Karsten Jürchott, Norbert Mages, and the Sequencing Facility of the Max-Planck-Institute for Molecular Genetics, Berlin, for their support in single-cell sequencing. We thank all patients and healthy donors for their participation.

This work was funded by the German Society of Rheumatology and Clinical Immunology (Forschungsinititative 2025 to PN), by the German Research Foundation under Germany’s Excellence Strategy (EXC-3105/1 – 533765739 to UW and UK) and by the German Research Foundation Individual Grants (WA 2765/5 and WA 2765/13 to UW).

## Literature

1. Gravallese EM, Firestein GS. Rheumatoid Arthritis — Common Origins, Divergent Mechanisms. New England Journal of Medicine. 2023;388(6):529–542. doi:10.1056/NEJMra2103726

2. Gregersen PK, Silver J, Winchester RJ. The shared epitope hypothesis. an approach to understanding the molecular genetics of susceptibility to rheumatoid arthritis. Arthritis & Rheumatism. 1987;30(11):1205–1213. doi:10.1002/art.1780301102

3. Eyre S, Bowes J, Diogo D, et al. High-density genetic mapping identifies new susceptibility loci for rheumatoid arthritis. Nat Genet. 2012;44(12):1336–1340. doi:10.1038/ng.2462

4. Wagner UG, Koetz K, Weyand CM, Goronzy JJ. Perturbation of the T cell repertoire in rheumatoid arthritis. Proc Natl Acad Sci USA. 1998;95(24):14447–14452. doi:10.1073/pnas.95.24.14447

5. Khan N, Shariff N, Cobbold M, et al. Cytomegalovirus Seropositivity Drives the CD8 T Cell Repertoire Toward Greater Clonality in Healthy Elderly Individuals. The Journal of Immunology. 2002;169(4):1984–1992. doi:10.4049/jimmunol.169.4.1984

6. Hashimoto K, Kouno T, Ikawa T, et al. Single-cell transcriptomics reveals expansion of cytotoxic CD4 T cells in supercentenarians. Proceedings of the National Academy of Sciences. 2019;116(48):24242–24251. doi:10.1073/pnas.1907883116

7. Pereira BI, De Maeyer RPH, Covre LP, et al. Sestrins induce natural killer function in senescent-like CD8+ T cells. Nat Immunol. 2020;21(6):684–694. doi:10.1038/s41590-020-0643-3

8. Brenchley JM, Karandikar NJ, Betts MR, et al. Expression of CD57 defines replicative senescence and antigen-induced apoptotic death of CD8+ T cells. Blood. 2003;101(7):2711–2720. doi:10.1182/blood-2002-07-2103

9. ElTanbouly MA, Noelle RJ. Rethinking peripheral T cell tolerance: checkpoints across a T cell’s journey. Nat Rev Immunol. Published online October 19, 2020. doi:10.1038/s41577-020-00454-2

10. Weyand CM, Goronzy JJ. Immune Aging in Rheumatoid Arthritis. Arthritis & Rheumatology. Published online February 11, 2025:art.43105. doi:10.1002/art.43105

11. Martens PB, Goronzy JJ, Schaid D, Weyand CM. Expansion of unusual CD4+ T cells in severe rheumatoid arthritis. Arthritis & Rheumatism. 1997;40(6):1106–1114. doi:10.1002/art.1780400615

12. Schmidt D, Goronzy JJ, Weyand CM. CD4+ CD7-CD28-T cells are expanded in rheumatoid arthritis and are characterized by autoreactivity. May 1, 1996. doi:10.1172/JCI118638

13. Li Y, Shen Y, Hohensinner P, et al. Deficient Activity of the Nuclease MRE11A Induces T Cell Aging and Promotes Arthritogenic Effector Functions in Patients with Rheumatoid Arthritis. Immunity. 2016;45(4):903–916. doi:10.1016/j.immuni.2016.09.013

14. Beck F, Nguyen P, Hoffmann A, et al. CD4+ CD8ALOW T Cell Clonal Expansion Dependent on Costimulation in Patients With Rheumatoid Arthritis. Arthritis & Rheumatology. 2024;76(12):1719–1729. doi:10.1002/art.42960

15. Miron M, Meng W, Rosenfeld AM, et al. Maintenance of the human memory T cell repertoire by subset and tissue site. Genome Med. 2021;13(1):100. doi:10.1186/s13073-021-00918-7

16. Danke NA, Koelle DM, Yee C, Beheray S, Kwok WW. Autoreactive T Cells in Healthy Individuals. The Journal of Immunology. 2004;172(10):5967–5972. doi:10.4049/jimmunol.172.10.5967

17. Wendeler MW, Paccaud J, Hauri H. Role of Sec24 isoforms in selective export of membrane proteins from the endoplasmic reticulum. EMBO Reports. 2007;8(3):258–264. doi:10.1038/sj.embor.7400893

18. Larbi A, Fulop T. From “truly naïve” to “exhausted senescent” T cells: When markers predict functionality. Cytometry Part A. 2014;85(1):25–35. doi:10.1002/cyto.a.22351

19. Fritsch RD, Shen X, Sims GP, Hathcock KS, Hodes RJ, Lipsky PE. Stepwise Differentiation of CD4 Memory T Cells Defined by Expression of CCR7 and CD27. The Journal of Immunology. 2005;175(10):6489–6497. doi:10.4049/jimmunol.175.10.6489

20. Rao DA, Gurish MF, Marshall JL, et al. Pathologically expanded peripheral T helper cell subset drives B cells in rheumatoid arthritis. Nature. 2017;542(7639):110–114. doi:10.1038/nature20810

21. Dunlap G, Wagner A, Meednu N, et al. Clonal associations between lymphocyte subsets and functional states in rheumatoid arthritis synovium. Nat Commun. 2024;15(1):4991. doi:10.1038/s41467-024-49186-0

22. Turcinov S, af Klint E, Van Schoubroeck B, et al. Diversity and Clonality of T Cell Receptor Repertoire and Antigen Specificities in Small Joints of Early Rheumatoid Arthritis. Arthritis & Rheumatology. 2023;75(5):673–684. doi:10.1002/art.42407

23. Venken K, Jarlborg M, Stevenaert F, et al. Shared lung and joint T cell repertoire in early rheumatoid arthritis driven by cigarette smoking. Annals of the Rheumatic Diseases. 2025;84(3):409–420. doi:10.1136/ard-2024-226284

24. Goëb V, Aegerter P, Parmar R, et al. Progression to rheumatoid arthritis in early inflammatory arthritis is associated with low IL-7 serum levels. Annals of the Rheumatic Diseases. 2013;72(6):1032–1036. doi:10.1136/annrheumdis-2012-202377

25. Wagner U, Schatz A, Baerwald C, Rossol M. Brief Report: Deficient Thymic Output in Rheumatoid Arthritis Despite Abundance of Prethymic Progenitors. Arthritis & Rheumatism. 2013;65(10):2567–2572. doi:10.1002/art.38058

26. Pierer M, Rossol M, Kaltenhäuser S, et al. Clonal expansions in selected TCR BV families of rheumatoid arthritis patients are reduced by treatment with the TNFα inhibitors etanercept and infliximab. Rheumatol Int. 2011;31(8):1023–1029. doi:10.1007/s00296-010-1402-9

27. Williams L, Saber AO, Awad S, et al. Divergent Effects of Cytomegalovirus and Rheumatoid Arthritis on Senescent CD4+ T Cells. Eur J Immunol. 2025;55(11):e70093. doi:10.1002/eji.70093

28. Baker KF, McDonald D, Hulme G, et al. Single-cell insights into immune dysregulation in rheumatoid arthritis flare versus drug-free remission. Nat Commun. 2024;15(1):1063. doi:10.1038/s41467-024-45213-2

29. Súkeníková L, Mallone A, Schreiner B, et al. Autoreactive T cells target peripheral nerves in Guillain–Barré syndrome. Nature. 2024;626(7997):7997. doi:10.1038/s41586-023-06916-6

30. Mitchell JS, Spanier JA, Dwyer AJ, et al. CD4+ T cells reactive to a hybrid peptide from insulin-chromogranin A adopt a distinct effector fate and are pathogenic in autoimmune diabetes. Immunity. 2024;57(10):2399–2415.e8. doi:10.1016/j.immuni.2024.07.024

31. Nanki T, Imai T, Nagasaka K, et al. Migration of CX3CR1-positive T cells producing type 1 cytokines and cytotoxic molecules into the synovium of patients with rheumatoid arthritis. Arthritis & Rheumatism. 2002;46(11):2878–2883. doi:10.1002/art.10622

32. Sawai H, Park YW, He X, Goronzy JJ, Weyand CM. Fractalkine mediates T cell– dependent proliferation of synovial fibroblasts in rheumatoid arthritis. Arthritis & Rheumatism. 2007;56(10):3215–3225. doi:10.1002/art.22919

33. Higashioka K, Sowerby JM, Marks KE, et al. PD-1 signalling restrains pathogenic T peripheral helper and effector CD4+ T cell functions in rheumatoid arthritis. Annals of the Rheumatic Diseases. Published online July 2026:S0003496726003559. doi:10.1016/j.ard.2026.06.007

34. Lê S, Josse J, Husson F. FactoMineR: An R Package for Multivariate Analysis. J Stat Soft. 2008;25(1). doi:10.18637/jss.v025.i01

35. Yang Q, Safina KR, Nguyen KDQ, Tuong ZK, Borcherding N. scRepertoire 2: Enhanced and efficient toolkit for single-cell immune profiling. Shinde P, ed. PLoS Comput Biol. 2025;21(6):e1012760. doi:10.1371/journal.pcbi.1012760

36. Hao Y, Stuart T, Kowalski MH, et al. Dictionary learning for integrative, multimodal and scalable single-cell analysis. Nat Biotechnol. 2024;42(2):293–304. doi:10.1038/s41587-023-01767-y

37. Yu G, Wang LG, Han Y, He QY. clusterProfiler: an R Package for Comparing Biological Themes Among Gene Clusters. OMICS: A Journal of Integrative Biology. 2012;16(5):284–287. doi:10.1089/omi.2011.0118

