## Supplementary table S3-S5 for "Hyperexpanded CD4^+^ T cell clones in rheumatoid arthritis show attenuated senescence and accumulate in afflicted joints"

**Table S3A. Patient characteristics — Validation cohort**

| **Characteristic** | **RA (n = 44)** | **HD (n = 44)** | **p value** |
| --- | --- | --- | --- |
| ***Demographics*** | | | |
| Age, years | 54.3 ± 10.5 | 53.8 ± 10.8 | 0.98 |
| Female sex, n (%) | 33 (75) | 22 (50) | **0.03* |
| ***Disease characteristics*** | | | |
| Disease duration, months | 60.2 ± 76.8 | — | — |
| RF positive, n (%) | 40 (90.1) | — | — |
| Anti-CCP positive, n (%) | 43 (97.7) | — | — |
| DAS28-CRP | 3.7 ± 1.4 | — | — |
| SDAI | 19.5 ± 13.3 | — | — |
| CRP | 13.0 ± 19.2 | — | — |
| ***Flow cytometry pilot cohort (n = 15 each)*** | | | |
| Age, years | 55.9 ± 9.7 | 54.8 ± 9.0 | 0.81 |
| Female sex, n (%) | 7 (46.7) | 10 (66.7) | 0.46 |
| Disease duration, months | 22 ± 35 | — | — |
| DAS28-CRP | 3.9 ± 1 | — | — |
| CMV-IgG positive, n (%) | 8 (53) | 9 (60) | >0.99 |

*Values are mean ± SD unless otherwise stated. HD, healthy donor; RA, rheumatoid arthritis; RF, rheumatoid factor; anti-CCP, anti-cyclic citrullinated peptide antibody; DAS28-CRP, Disease Activity Score in 28 joints with C-reactive protein; SDAI, Simple Disease Activity Index; CRP, C-reactive protein. p values: Mann–Whitney U test (continuous) or Fisher's exact test (categorical). — indicates not applicable.*

**Table S3B. Patient characteristics — Synovial cohort**

| **Characteristic** | **RA blood (n = 35)** | **RA syn. tissue (n = 18)** | **RA syn. fluid (n = 20)** |
| --- | --- | --- | --- |
| ***Demographics*** | | | |
| Age, years | 60.2 ± 16.8 | 64.9 ± 13.1 | 62.3 ± 16.8 |
| Female sex, n (%) | 28 (80) | 4 (24) | 2 (11) |
| ***Disease characteristics (RA)*** | | | |
| Disease duration, years | 11.3 ± 9.4 | 13.5 ± 8.1 | 9.8 ± 9.6 |
| DAS28-CRP | 4.3 ± 1.1 | 4.1 ± 1.2 | 4.6 ± 1.3 |
| RF positive, n (%) | 25 (71) | 9 (75) | 10 (71) |
| Anti-CCP positive, n (%) | 20 (57) | 8 (67) | 8 (62) |
| ***Current treatment (RA), n (%)*** | | | |
| Glucocorticoids only | 5 (15) | 1 (6) | 4 (22) |
| csDMARD ± glucocorticoids | 10 (30) | 5 (29) | 4 (22) |
| bDMARD / tsDMARD ± glucocorticoids ± csDMARD | 10 (30) | 7 (41) | 4 (22) |
| ***Synovial joint, n (%)*** | | | |
| Knee | — | 6 (35) | 11 (61) |
| Hip | — | 3 (18) | — |
| Wrist | — | 3 (18) | 1 (6) |
| Elbow | — | 2 (12) | 2 (11) |
| Small joints | — | 1 (6) | 2 (11) |
| Ankle | — | 1 (6) | — |
| Shoulder | — | — | 1 (6) |

*Values are mean ± SD unless otherwise stated. syn., synovial. csDMARD, conventional synthetic disease-modifying anti-rheumatic drugs; b/tsDMARD, biological or targeted synthetic disease-modifying anti-rheumatic drugs. — indicates not applicable.*

**Table S3C. Patient characteristics — Abatacept cohort**

| **Characteristic** | **RA (n = 6)** |
| --- | --- |
| ***Demographics*** | |
| Age, years | 56.7 ± 10.7 |
| Female sex, n (%) | 4 (66.7) |
| ***Disease characteristics*** | |
| Disease duration, years | 2.3 ± 2.6 |
| DAS28-CRP at baseline | 4.06 ± 0.85 |
| SDAI at baseline | 20.0 ± 7.3 |
| DAS28-CRP at week 12 | 2.62 ± 0.70 |
| SDAI at week 12 | 9.6 ± 4.7 |
| RF positive, n (%) | 6 (100) |
| Anti-CCP positive, n (%) | 6 (100) |

*Values are mean ± SD unless otherwise stated.*

**Table S4. Multivariable linear regression**

Dependent variable: frequency of CCR7^-^CD27^-^ among all CD4^+^ T cells in synovial tissue

Predictors: DAS28-CRP, Age (years)

Cases: n = 15

Goodness of fit: R^2^ = 0.6578

| **Parameter estimates** | **Variable** | **Estimate** | **95% CI (profile likelihood)** | **P value** |
| --- | --- | --- | --- | --- |
| β1 | DAS28-CRP | 4,227 | 0,8157 to 7,638 | 0,0193 |
| β2 | Age | 0,5597 | 0,2431 to 0,8762 | 0,0023 |

**Table S5. Flow cytometry antibody panel**

| **Marker** | **Clone** | **Fluorochrome** | **Vendor** | **Dilution** |
| --- | --- | --- | --- | --- |
| Viability dye | — | BV510 (405/520) | Miltenyi Biotec | 1:200 |
| CD3 | REA613 | PerCP-Vio® 700 | Miltenyi Biotec | 1:100 |
| CD4 | M-T321 | APC-Vio® 770 | Miltenyi Biotec | 1:50 |
| CD8α | REA734 | FITC | Miltenyi Biotec | 1:100 |
| CD8β | 2ST8.5H7 | BUV395 | BD Biosciences | 1:100 |
| CD27 | L128 | BUV737 | BD Biosciences | 1:20 |
| CCR7 | 150503 | BV421 | BD Biosciences | 1:20 |
| PD-1 (CD279) | REAfinity™ | Vio® Bright B515 | Miltenyi Biotec | 1:20 |
| CD25 | 3G10 | APC | Miltenyi Biotec | 1:20 |
| Granzyme B (GZMB) | REA226 | PE | Miltenyi Biotec | 1:100 |
| Perforin (PRF1) | REA1061 | APC | Miltenyi Biotec | 1:100 |
| IL-7R (CD127) | REA614 | PE | Miltenyi Biotec | 1:20 |
| IL-6Rα (CD126) | REA291 | APC | Miltenyi Biotec | 1:10 |
| CX3CR1 | REA385 | PE | Miltenyi Biotec | 1:100 |
| CD62L (SELL) | 145/15 | APC | Miltenyi Biotec | 1:50 |
| HLA-DR | REA805 | PE | Miltenyi Biotec | 1:50 |
| CD25 (IL2RA) | 3G10 | APC | Miltenyi Biotec | 1:50 |
| CD28 | REA612 | APC | Miltenyi Biotec | 1:50 |
| GPR56 (ADGRG1) | REA467 | APC | Miltenyi Biotec | 1:50 |
| SLAMF7 | REA150 | PE | Miltenyi Biotec | 1:100 |
| KLRG1 | REA261 | PE-Vio® 615 | Miltenyi Biotec | 1:50 |
| KLRF1 (NKp80) | REA845 | APC ᶜ | Miltenyi Biotec | 1:50 |
| NKG2D (KLRK1) | REA797 | PE | Miltenyi Biotec | 1:50 |
| CD56 (NCAM1) | AF12-7H3 | APC | Miltenyi Biotec | 1:50 |

*All Miltenyi REAfinity clones are recombinant human IgG1. Clone M-T321 (CD4), AF12-7H3 (CD56), 145/15 (CD62L), and 3G10 (CD25) are mouse IgG1κ. BD clones L128 (CD27) and 150503 (CCR7) are mouse IgG1/IgG2α; 2ST8.5H7 (CD8β) is mouse IgG2ακ.*
