## Supplementary figures for "Hyperexpanded CD4^+^ T cell clones in rheumatoid arthritis show attenuated senescence and accumulate in afflicted joints"

**Supplementary material**

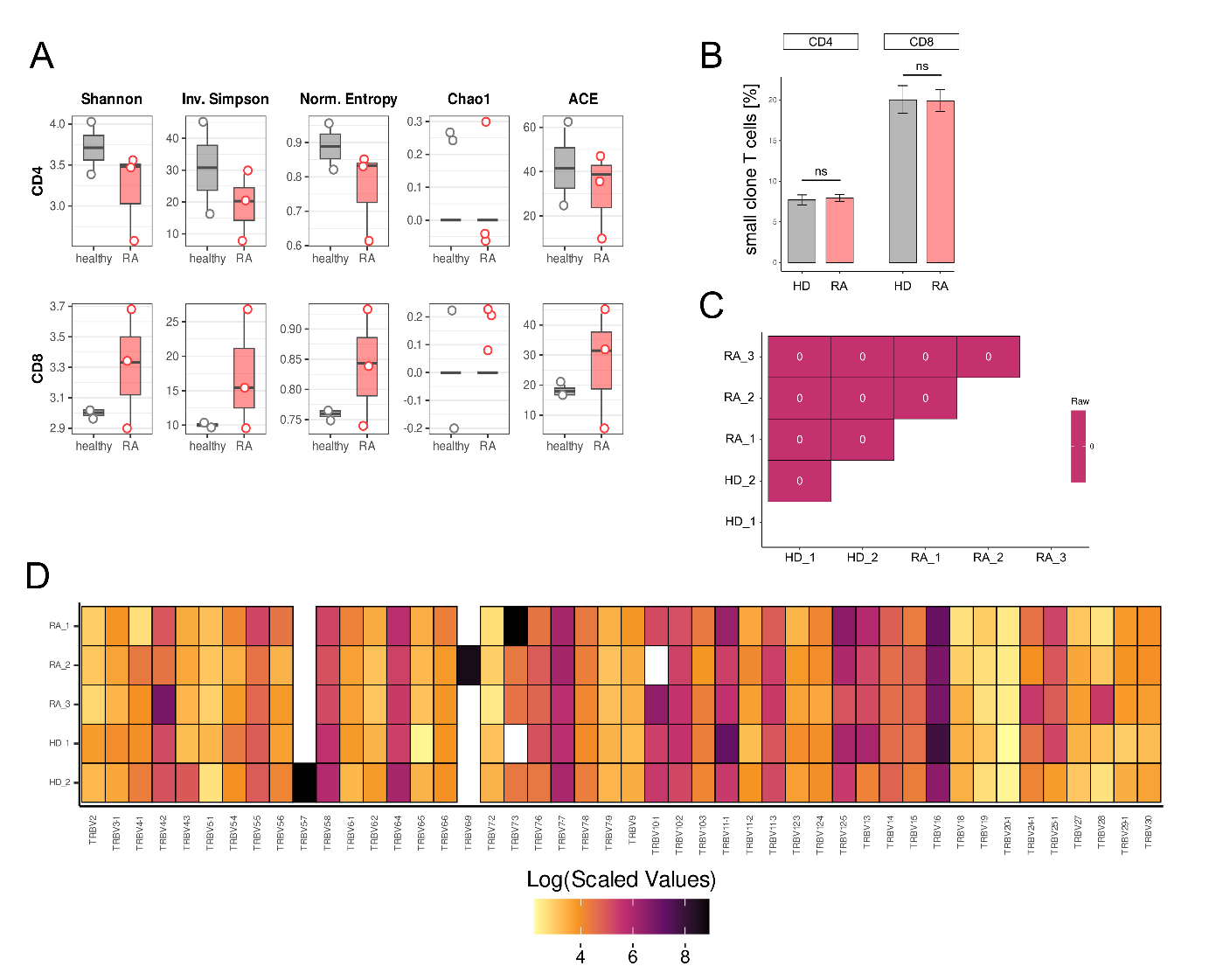
**Figure S1: Clonality analysis of the single-cell TCR data.**

**(A**) Boxplots displaying diversity and clonality indices of clonally expanded CD4⁺ and CD8⁺ T cells in RA and HD. **(B)** Proportion of T cells belonging to a small, conventional CD4^+^ or CD8^+^ clone amongst all CD4^+^ or CD8^+^ T cells did not statistically differ between RA and HD. Small clones are defined as comprising >1 and <10 T cells with identical paired α/β-TCR sequence. **(C)** No clonal overlap between the samples. Clones are defined as cells sharing the same paired α/β-TCR sequence. **(D)** Gene usage of the TCR β-chain variable (V) region of each individual.

Inv. Simpson, Inverse Simpson index; Norm. Entropy, Normalized Entropy index; ACE, Abundance-based Coverage Estimator. TCR, T cell receptor.

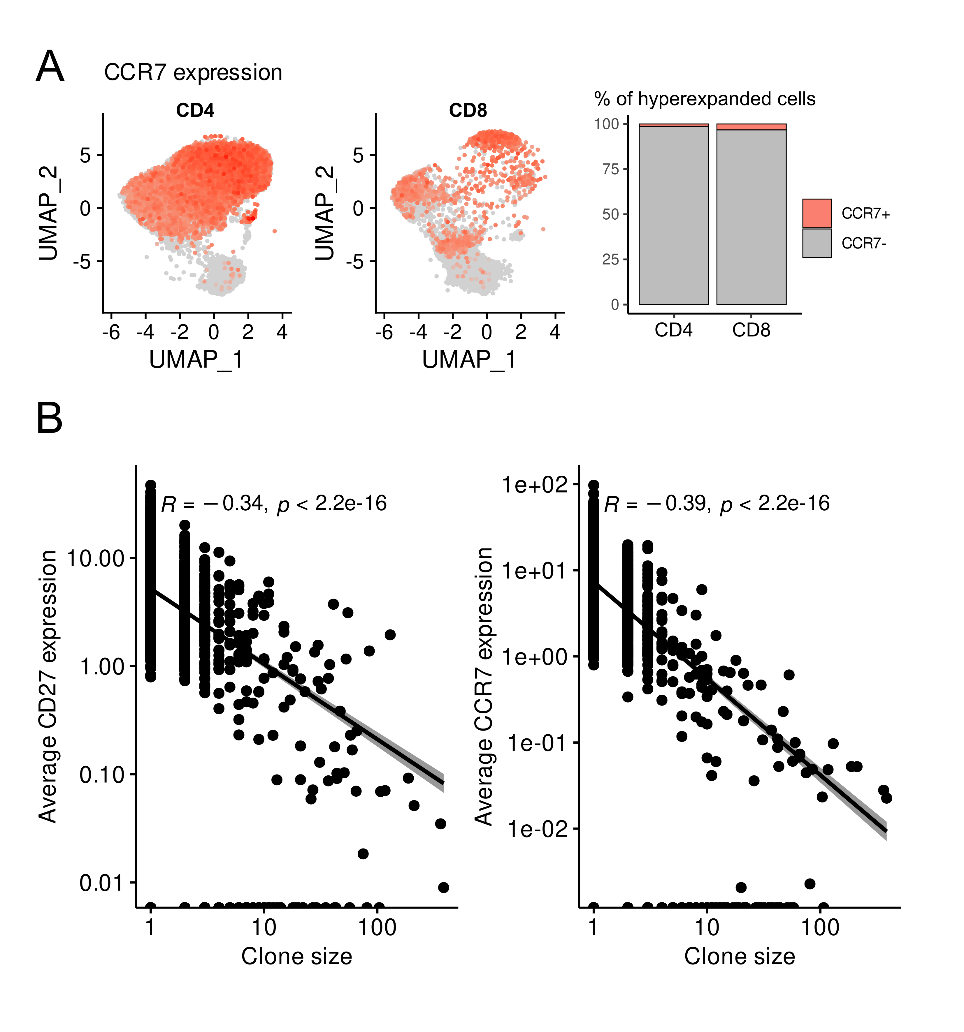

**Figure S2: CCR7 and CD27 loss is associated with clonal expansion.**

**(A**) UMAP showing CCR7 expression for CD4^+^ and CD8^+^ T cells (left and middle panel), alongside the proportion of cells expressing CCR7 in hyperexpanded T cell clones (right panel). **(B)** Spearman’s correlation between clone size and average expression of CCR7 and CD27 in each clone.

UMAP, Uniform Manifold Approximation and Projection.

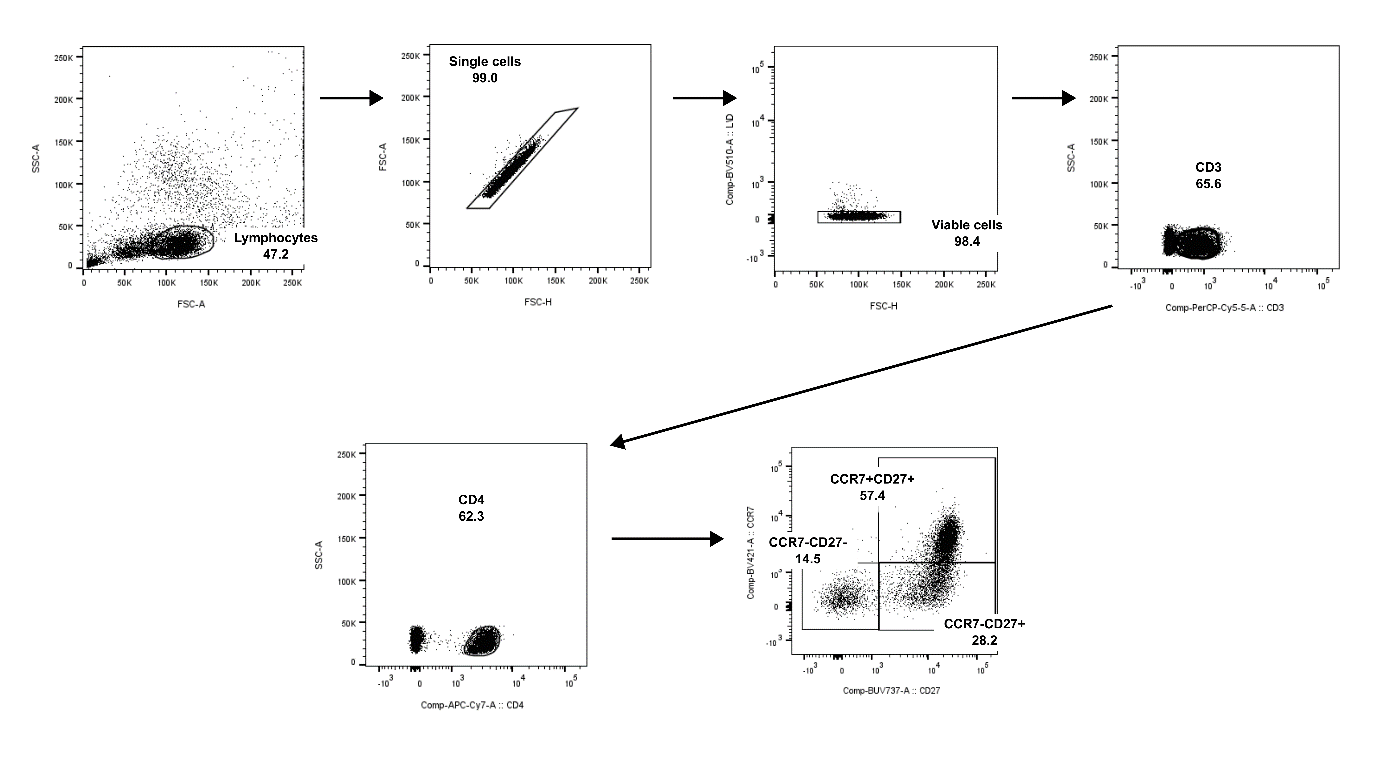
**Figure S3: Flow cytometry analysis of senescence-associated markers.**

Gating strategy for the identification of CCR7^-^CD27^-^CD4^+^ T cells.

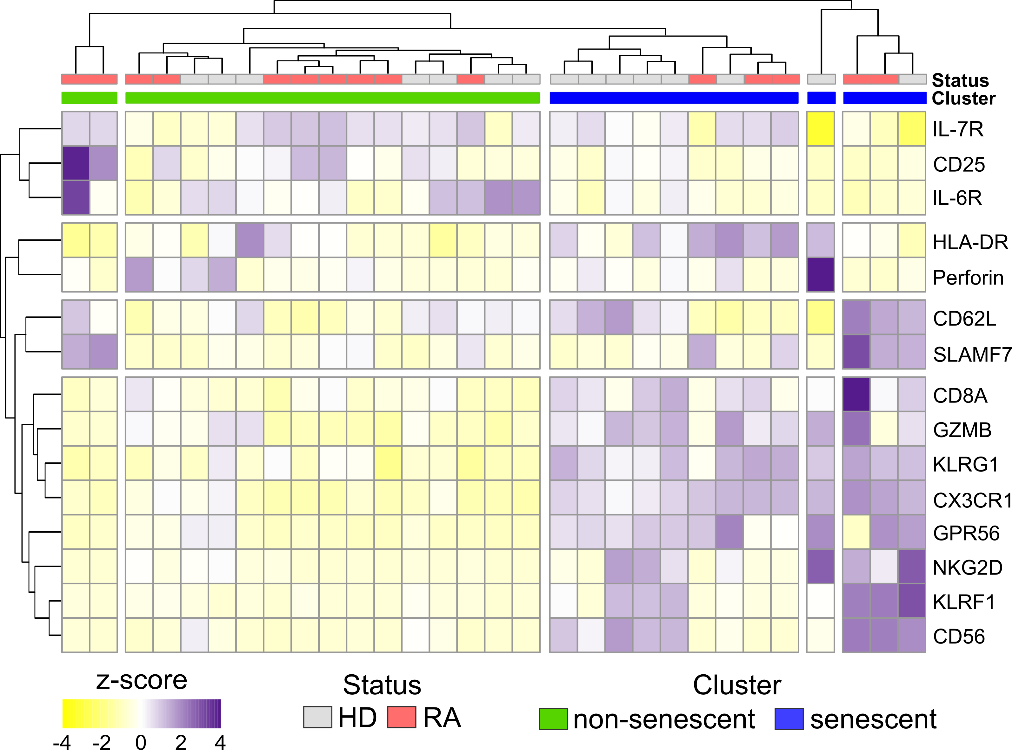

**Figure S4: Hierarchical clustering of marker expression in hyperexpanded CD4 T cell clones.** Hierarchical clustering resulted in two non-senescent clusters (low expression of NK and cytotoxicity markers, high expression of CD25, IL7- and IL-6 receptors) and three senescent clusters (high expression of NK and cytotoxicity markers). RA samples are overrepresented in non-senescent clusters, HD samples in senescent clusters (n=15 each).

NK, natural killer; RA, rheumatoid arthritis; HD, healthy donors.

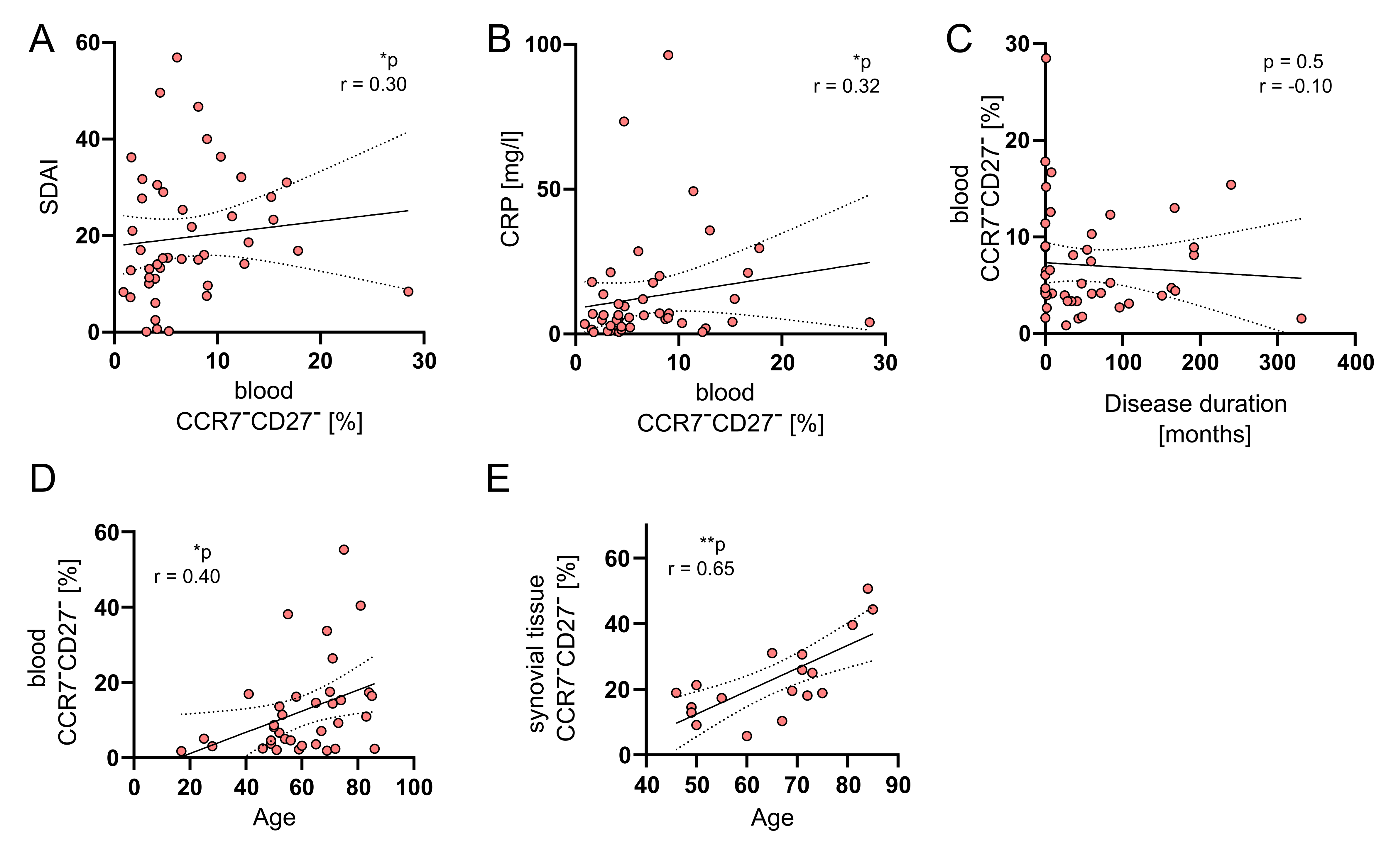

**Figure S5:** Frequency of hyperexpanded CD4^+^ clones in RA peripheral blood (n=45) correlate with **(A)** Simplified Disease Activity Index (SDAI) and **(B)** C-reactive protein (CRP), but not with **(C)** disease duration. Correlation of age with hyperexpanded CD4^+^ clone frequency in RA **(D)** peripheral blood (n=35) and **(E)** synovial tissue (n=20).

RA, rheumatoid arthritis.
